# Modelling infectious disease transmission potential as a function of human behaviour

**DOI:** 10.1101/2021.07.16.21260521

**Authors:** Caroline E. Walters, Maria Bekker-Nielsen Dunbar, Dale Weston, Ian M. Hall

## Abstract

Mathematical modelling is an important public health tool for aiding understanding the spread of respiratory infectious diseases, such as influenza or COVID-19, and for quantifying the effects of behavioural interventions. However, such models rarely explicitly appeals to theories of human behaviour to justify model assumptions. Here we propose a novel mathematical model of disease transmission via fomites (luggage trays) at airport security screening during an outbreak. Our model incorporates the self-protective behaviour of using hand sanitiser gel in line with the extended parallel processing model (EPPM) of behaviour.

We find that changing model assumptions of human behaviour in line with the EPPM gives qualitatively different results on the optimal placement of hand sanitiser gels within an airport compared to the model with naive behavioural assumptions. Specifically, that it is preferable to place hand sanitiser gels after luggage screening in most scenarios, however in situations where individuals perceive high threat and low efficacy this strategy may need to be reviewed. This model demonstrates how existing behavioural theories can be incorporated into mathematical models of infectious disease.

## 1 Introduction

Mathematical models for the spread of respiratory infectious diseases provide important public health insights to policymakers regarding non-pharmaceutical interventions (e.g. hand hygiene, mask wearing, social distancing), most recently during the COVID-19 pandemic. Behaviour of individuals is a key driver of epidemic disease dynamics (Ferguson, 2007; Durham et al., 2012; Weston et al., 2018; Atchison et al., 2021; West et al., 2020), however behavioural theories are rarely mentioned within epidemiological model descriptions (Weston et al., 2018). As both behavioural science and epidemiological modelling disciplines feed in to advising policymakers during outbreak (e.g. Scientific Advisory Group for Emergencies in the UK (SAGE)), we posit that incorporating specific behavioural theories into the mathematical models will enable an easier translation of modelling results into practical guidance for the population and the ability to test how different behavioural assumptions affect optimal health-protective strategies.

Here we present a novel model for indirect contact transmission of a pandemic respiratory virus within an airport setting, where population compliance with a hand hygiene intervention is informed by a specific behavioural science theory. Models of self-inoculation through hand contamination exist (Nicas and Best, 2008; Pham et al., 2020; Teslya et al., 2020) but are not constructed from a behavioural science perspective. We selected the Extended Parallel Processing Model (EPPM), proposed by Witte (1992), over a vast array of potential behaviour change theories (see Michie et al. (2014)) as the basis for the behavioural component of our model for several reasons. First, it is one of very few behaviour change theories tailored specifically to infectious disease response. Secondly, very few behaviour change theories explicitly model emotions within the decision making process, and none of the gold standard papers incorporated in Weston et al. (2018) explicitly model emotional responses or address the EPPM specifically. Third, perceptions regarding the H1N1 influenza pandemic were found to be predictors of compliance with recommended health-protective behaviours (Rubin et al., 2009, 2010), with figures on hand hygiene during the 2009 H1N1 influenza pandemic, a milder pandemic than COVID-19, showing only moderate compliance with interventions (Pandemic Influenza Preparedness Team, 2011). Finally, the EPPM framework has previously been used to examine hand hygiene (Botta et al., 2008).

EPPM purports that individuals engage in two appraisal processes in response to a health related message: individuals appraise a health related message in terms of the threat that it poses to the self. If this threat is sufficiently high, the individual will experience fear, which stimulates an appraisal concerning an individual’s perceived ability to deal with the threat (perceived efficacy). Message acceptance (and adoption of recommended behaviours) is a function of these threat and efficacy appraisals: when both threat and efficacy are high, i.e. an individual recognises danger and feels able to deal with it, individuals respond to the danger and not to their fear, and so are likely to accept the message and behave adaptively. However, when high threat is matched with low efficacy, individuals experience intensified fear and so engage in maladaptive, defensive responses where they ignore the message or recommended behaviour in order to alleviate their fear.

We consider the chance of indirect contact via fomite transmission from airport security luggage screening trays motivated by the results of Ikonen et al. (2018). They sampled surfaces in passenger areas of Helsinki airport and detected respiratory virus nucleic acid on the luggage trays, which they highlight as a potential disease transmission risk. Influenza viruses can survive on certain non-porous surfaces, such as plastic and metals, for 24-48 hours (Weber and Stilianakis, 2008) and SARS-CoV-2 for up to 72 hours (van Doremalen et al., 2020). Fomite transmission of respiratory pathogens may occur (World Health Organization, 2020; Zhou et al., 2020), although the contribution to transmission in real world settings, rather than controlled laboratory settings, is uncertain (see Boone and Gerba, 2007; Goldman, 2020; Otter et al., 2016; Weber and Stilianakis, 2008, for more in-depth discussion). Despite high virus inactivation rates on human hands, individuals frequently touch their face, for example eyes or mouth, which are ‘portals of entry’ (Boone and Gerba, 2007) therefore fomite transmission may be a viable transmission route if high doses of virus persist on frequently-touched non-porous surfaces (Weber and Stilianakis, 2008). As available evidence is not conclusive on the role of fomite transmission, we consider the contamination of an individual’s hands with a respiratory pathogen from touching non-porous surfaces to be a possible mechanism in virus transmission worthy of investigation.

An intervention to interrupt such transmission is good hand hygiene, either hand-washing with soap and water or using of hand sanitiser gels. Human coronaviruses, including SARS-CoV-2, are inactivated by hand sanitiser gel (Kratzel et al., 2020; Kampf et al., 2020) and Larson et al. (2012) found that hand sanitiser foam, gel and wipes all significantly reduced viral load on hands. Warren-Gash et al. (2012) find that the efficacy of hand hygiene interventions on reducing transmission of respiratory viruses within community settings is dependent on multiple factors, related to the setting, context, and compliance. For instance, Cowling et al. (2009) find the combined use of facemasks and hand hygiene had a statistically significant effect on reducing influenza transmission in households under certain circumstances, however the effect of each individual intervention alone could not be determined; Godoy et al. (2012) found marginal benefits from the use of hand sanitiser gel in reducing hospitalisation from influenza. As each study (see also Aiello et al. (2008, 2010, 2012); Suess et al. (2012)) looks at different conditions, their results so cannot be directly compared, nor assumed to apply to, the airport setting. In the absence of any airport specific studies, we consider indirect contract via fomites to be a potential transmission route worthy of investigation.

## 2 Probabilistic model of disease contamination

We propose a probabilistic model for the chance of individual picking up virus on their hands after handling luggage trays at airport security. Individuals and luggage trays can be in one of two states: susceptible, where no virus is present; or contaminated, where virus is present on human hands or on the surface of the luggage trays. As individuals pass through security screening there is a chance of transference of a pathogen from either human to tray or tray to human. Individuals pass through the system only once, whereas trays are repeatedly handled by multiple individuals. Figure 1 show the decision process for each each individual in the model. When they first arrive at security screening they have probability *π* of being contaminated. During luggage screening they will then have the option to comply with a health intervention — the use of hand sanitiser gel. We assume that an individual uses available hand sanitiser gel before handling the luggage screening trays with probability *ρ* (compliance before) and uses available hand sanitiser gel after handling the luggage screening trays with probability *ω* (compliance after). We assume that using hand sanitiser gel is 100% effective in killing any virus present on hands and therefore use of hand sanitiser gel switches the status of contaminated humans to susceptible. Finally we assume that an individual becomes contaminated after handling the security trays with probability *α*. We are interested in the probability that an individual leaves the process contaminated as a function of their compliance. By considering all possible branches of the tree in Figure 1, we find that the overall probability of an individual leaving contaminated is given by

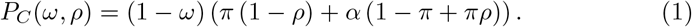

**Figure 1:**
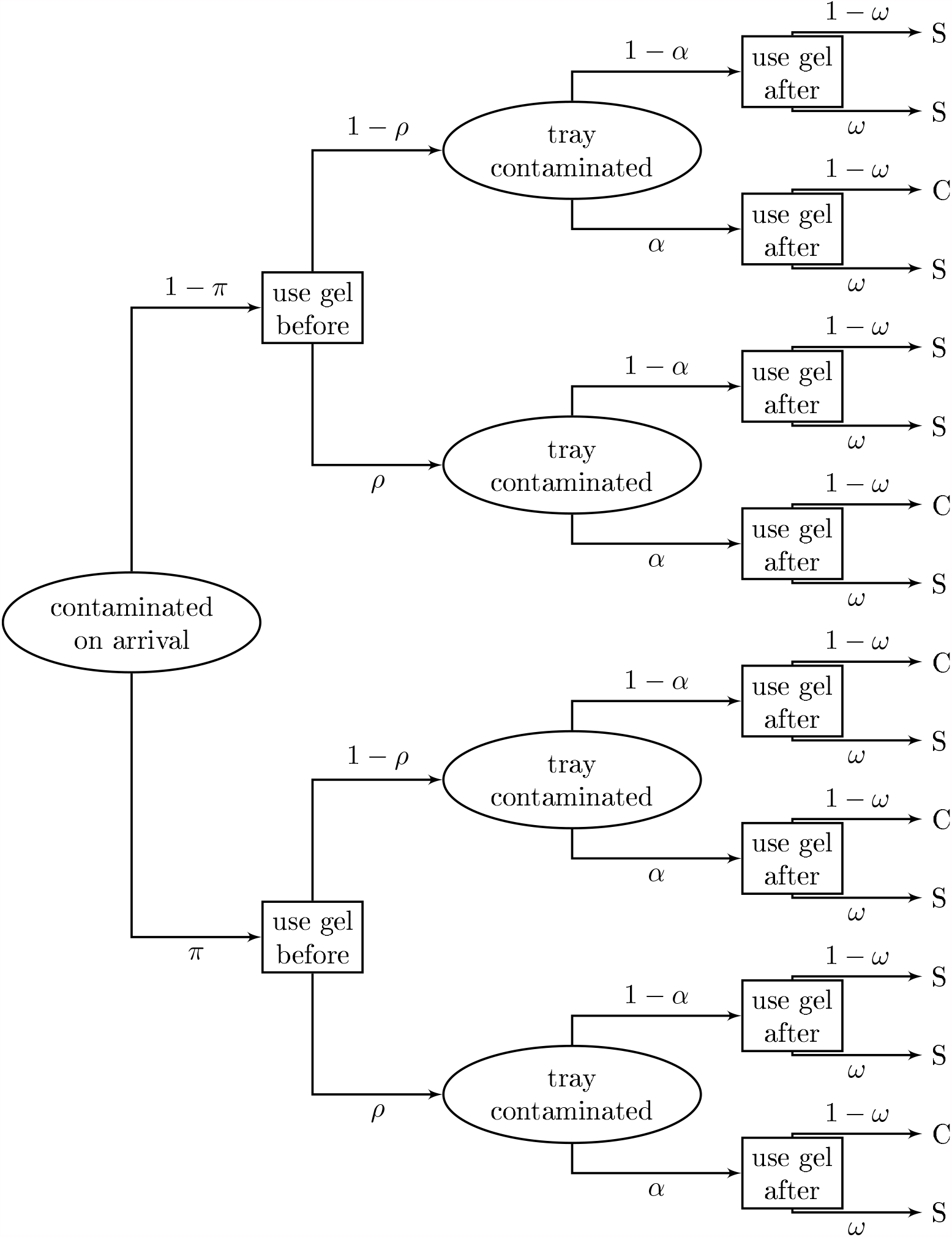
Decision tree showing progression through airport security luggage screening. Individuals leave the process either susceptible (S) or contaminated (C). Rectangles represent decision nodes, ovals are chance nodes.

The parameter *α* ∈ [0, 1] accounts for a single individual’s contact with trays. It comprises the probability of a tray being contaminated, Θ ∈ [0, 1], and the probability of a single contaminated tray contaminating a human, *γ* ∈ [0, 1]. For a single human contacting *m* ∈ ℕ trays, the probability of contamination transfer is

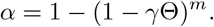

We assume the probability of a luggage screening tray contaminating a human to be small as we are not aware of this process having been highlighted as a major route for disease transmission in any previous outbreaks. Thus we assume *γ*Θ ≪ 1 and use a first order approximation to find

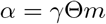

for instances of *m* > 1.

We assume that contamination of a tray occurs after it has been handled by a contaminated individual, thus the contamination status of trays will change over time. We model this using a Markov birth-death process with a population of humans, size *H*, and a population of trays, size *T*. The probability of a tray being contaminated is Θ = *T*_*C*_*/T*, where *T*_*C*_ is the number of contaminated trays. We assume the number of contaminated humans is *H*_*C*_ = *πH* as, once individuals have passed through security screening they then leave our model system. The probability that a tray becomes contaminated is given by the probability that the tray is susceptible and the tray has at least one interaction sufficient for contaminant transfer with a contaminated human. We define: *c*, the number of humans that a single tray contacts per hour; *β*, the probability that a contaminated human transmits to a tray; *T* − *T*_*C*_, the number of susceptible trays. The probability of contaminant transmission from a contaminated human to a tray is then 1 − (1 − *β*)^*c*^ = *βc* by first order approximation as we assume that the chance of contaminant transmission is small (*β* ≪ 1). Thus a tray becomes contaminated, *T*_*C*_ → *T*_*C*_ + 1, with rate (1 − *ρ*)*βcH*_*C*_(*T* − *T*_*C*_)*/H* and a tray loses its contamination, *T*_*C*_ → *T*_*C*_ − 1, with rate *δT*_*C*_ where *δ* is the decay rate of the pathogen on the tray’s surface.

We assume trays are not cleaned throughout the day and that enough time has passed for steady state behaviour to be applicable. At steady state, births and deaths are equal, so the steady state probability of a tray being contaminated is

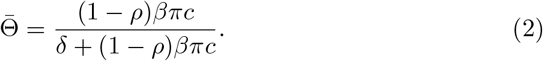

The probability of contamination, equation (1), at steady state becomes

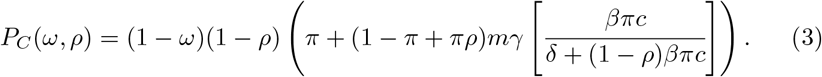

We are now interested in how the placement of the health intervention (hand sanitiser gel) affects the probability of contamination of an individual, *P*_*C*_.

### 2.1 Location of the intervention

From a baseline scenario of no intervention (*ρ* = 0 and *ω* = 0, equivalent to zero compliance with intervention), we define two natural measures of disease burden. First, the burden of infection contamination on trays themselves,

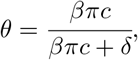

derived from the birth-death process (equation (2)) at steady state. Second, the additional burden of contamination in humans after contact with trays,

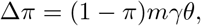

which is the difference between *P*_*C*_ (equation (3)) and the initial probability of contamination in humans, *π*. With no intervention available, equation (3) becomes

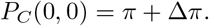

For the most general case (intervention available both before and after luggage screening) we can rewrite equation (3) as

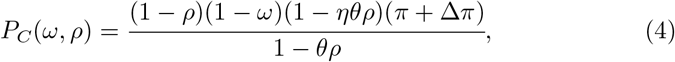

where

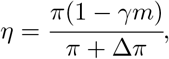

*η* ∈ (0, 1), and given *γ* is a probability expected to be less than 1*/m*. With appropriate substitution into equation (4), we find

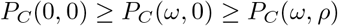

and

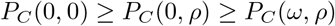

so, unsurprisingly, the probability of infection is highest if there is no intervention and lowest if there is an intervention both before and after luggage screening. If, however, hand sanitiser gel is available in only one location, either before or after luggage screening, then we wish to know which location minimises the probability of contamination. We investigate this by calculating the difference between both scenarios, *P*_*C*_(0, *ρ*) − *P*_*C*_(*ω*, 0), and divide by (*π* + Δ*π*) for convenience. This gives

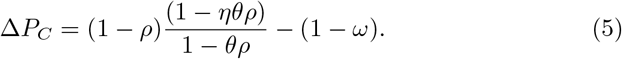

If Δ*P*_*C*_ *>* 0 then it is always optimal to place hand sanitiser gel after luggage screening to minimise the probability of contamination.

## 3 Modelling intervention compliance behaviour

In the first instance we assume that compliance is constant (*ρ* = *ω*), irrespective of intervention location. As

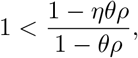

equation (5) shows that Δ*P*_*C*_ *>* 0 therefore the probability of infection is always reduced by placing the hand sanitiser gel intervention after luggage screening rather than before. We now choose to assume that compliance differs depending on the location of the hand sanitiser gel, informed by the EPPM behavioural theory.

As discussed in the introduction, EPPM is concerned with an individuals response to a health-protective message determining whether they engage with the recommended self-protective behaviour. Individuals engage in both threat and efficacy appraisal to determine their compliance with the proposed intervention. Here the message is to use hand sanitiser gel which has two public health benefits: 1) to prevent susceptible individuals from becoming contaminated; 2) to prevent infected individuals from contaminating the environment, which could then pose a risk to other individuals. In our example of placing hand sanitiser gel prior to luggage screening, individuals have not engaged in risky behaviour — the handling of trays — and therefore there is no threat to appraise. It is only after handling the trays that an individual has engaged in behaviour that could have negative health consequences for the individual, and thus EPPM could apply with threat appraisal taking place. For mathematical convenience, we assume that the threat and efficacy appraisal process does occur prior to luggage screening however assume that perceived threat prior to handling the trays is always less than the perceived threat after. Whilst this model assumption may not accurately represent behaviour in line with EPPM, we can interpret our model as having some baseline compliance prior to luggage screening that can be written in the same functional form as compliance after.

We define *ϵ* ∈ [0, 1] to be a measure of perceived efficacy and assume that it is the same either before and after luggage screening. We define *τ* ∈ [0, 1] to be a measure of perceived threat where *τ*_*before*_ *< τ*_*after*_ and introduce *k* ∈ [1, 1*/τ*] such that *τ*_*before*_ = *τ* and *τ*_*after*_ = *kτ*. We now define compliance as a function of threat and efficacy, *f* (*τ, ϵ*), such that compliance before and compliance after are given by

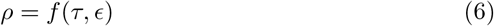

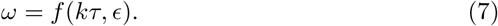

We now consider the form of the compliance function, *f* (*τ, ϵ*), in line with EPPM theory.

### 3.1 Increasing *f* (*τ, ϵ*) as *τ* and *ϵ* increase

If *f* (*τ, ϵ*) is increasing as *τ* and *ϵ* increase then, for any *k* > 1, we have *f* (*τ, ϵ*) < *f* (*kτ, ϵ*), hence *ρ* < *ω*. An example function can be seen in Figure 2. Equation (5) becomes

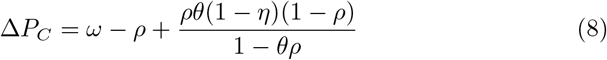

which has positive right hand side, therefore Δ*P*_*C*_ > 0 meaning it is always optimal to place hand sanitiser gel after luggage screening to minimise the probability of contamination.

**Figure 2:**
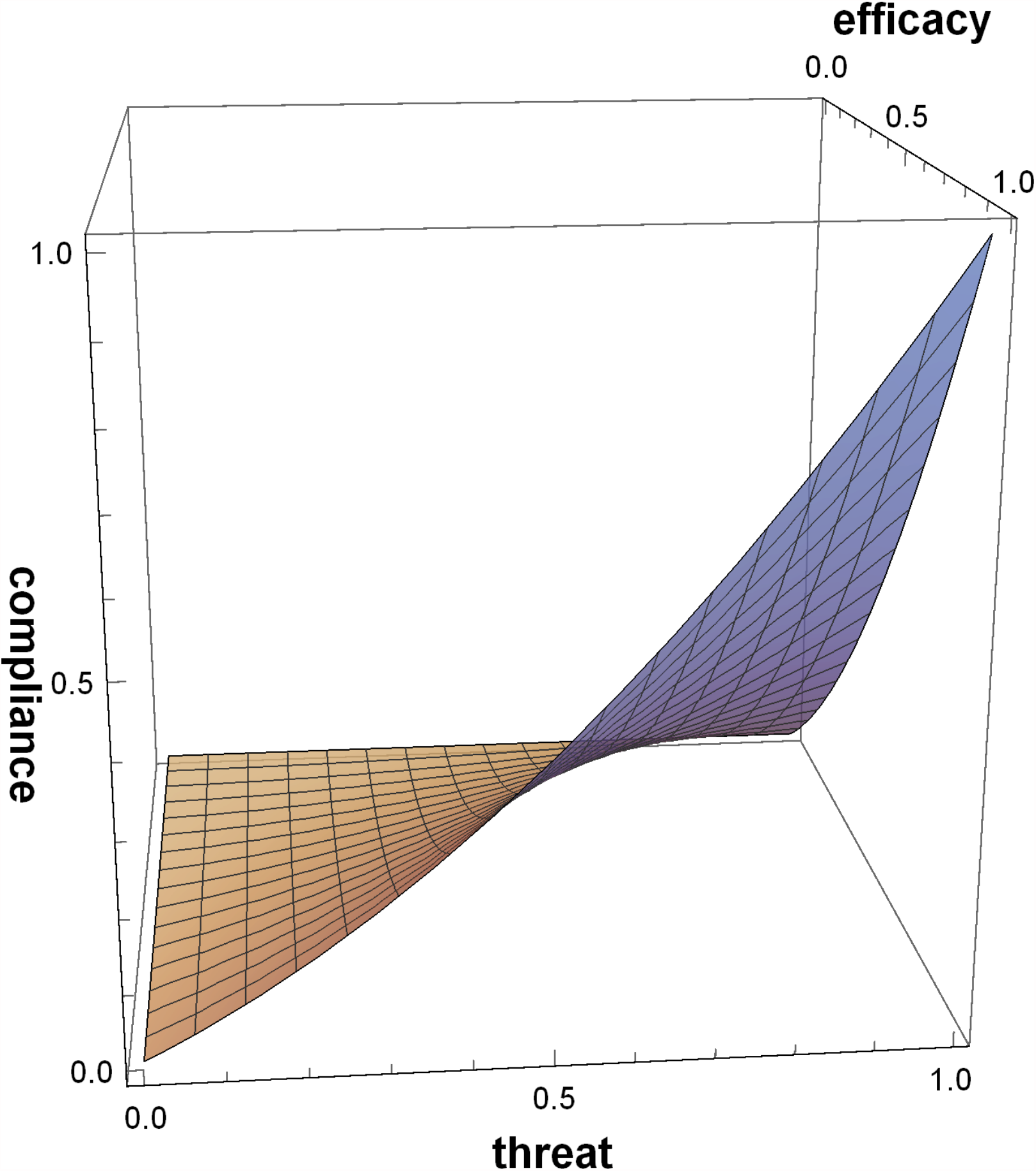
Compliance function which increases as both threat or efficacy increase.

### 3.2 Non-increasing *f* (*τ, ϵ*) as *τ and ϵ* increase

Now we consider the probability of compliance, aligned with EPPM theory, such that there is some threat threshold value beyond which the compliance function decreases with respect to threat. This formulation can represent individuals defensively avoiding or ignoring the health protective message because of having a high perceived threat combined with a low perceived efficacy in the health protective behaviour. We choose *f* (*τ, ϵ*) = *ϵτ* (*ϵ* + 1 − *τ*) which, substituting into equations (6) and (7), gives

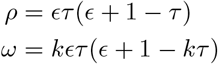

for *k* ∈ [1, 1/*τ*]. This means low threat, low efficacy gives low compliance; high threat, high efficacy gives high compliance; low threat, increasing efficacy gives increasing compliance; increasing threat with only low to middling efficacy gives some increase of compliance at first, but then switches to compliance decreasing with threat. This can be seen in Figure 3. In this final case, for a fixed threat, compliance should always increase with efficacy. Substituting into equation (5), we find

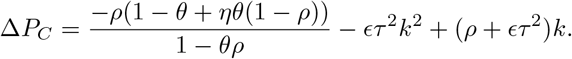

**Figure 3:**
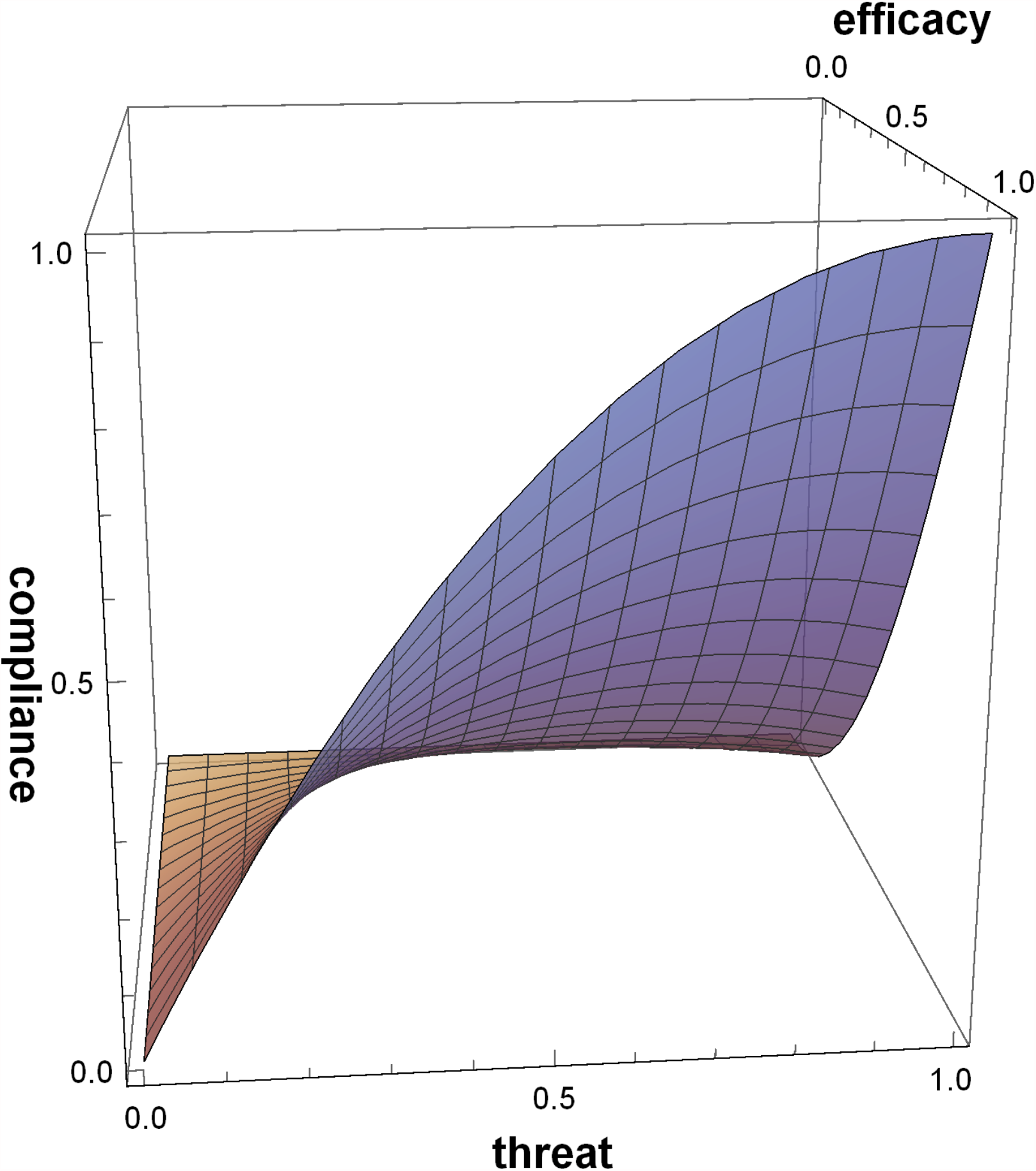
Compliance function which increases as efficacy increases but may increases then decreases as threat increases.

We can write this equation as a quadratic in *k*:

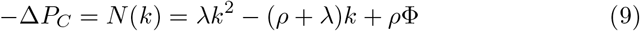

with positive coefficients

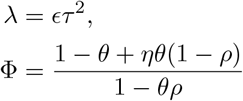

where *λ* ∈ [0, 1] and Φ ∈ [0, 1].

We now investigate if hand sanitiser gel before luggage screening is ever optimal. This corresponds to *N*(*k*) > 0, noting that *k* ∈ [1, 1/*τ*] by definition. As the leading coefficient of *N*(*k*) is positive by definition, *N*(*k*) has a minimum turning point. As *N*(1) = *ρ*(Φ – 1) ≤ 0 there must be a real root, *K*, such that *K* ≥ 1. If *K* < 1/*τ* then there exists a region in parameter space where *N*(*k*) > 0 for *k* ∈ [1, 1/*τ*] and thus conditions under which it is optimal to place hand sanitiser gel prior to luggage screening. We know

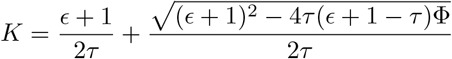

and we seek *K* < 1/*τ* which simplifies to

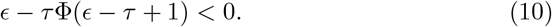

Rearranging inequality (10) we obtain an upper bound on *ϵ*

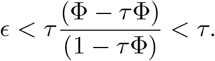

Therefore *ϵ* < *τ* is a necessary, but not sufficient, condition for hand sanitiser gel to be optimally placed prior to luggage screening.

For illustration we investigate how a change in disease prevalence (*π*) affects the region of threat-efficacy parameter space where placing hand sanitiser gel before luggage screening is optimal. We assume that each person uses a single tray, so *m* = 1. If there are 40 trays per queue and 10 queues at luggage screening then there are 400 trays in use each day. For 20,000 passengers per day, each tray is touched by 50 people, so *c* = 50. We also allow the virus to remain viable on the tray surface for 1 day (*δ* = 1). Choosing *β* = *γ* = 1*/*15, Figure 4 shows

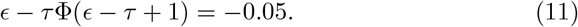

for different values of *π*. The critical region, where hand sanitiser gel before luggage screening is optimal, is the region below the curve and above the *τ* axis. As the value of *π* increases, corresponding to an increase in disease prevalence, the critical region initially diminishes in size. This fits with our EPPM behavioural assumptions where an initial increase in prevalence would lead to a greater chance of an individual perceiving a threat and thus engaging in efficacy appraisal, and adoption of the health protective behaviour (hand sanitiser gel use after handling luggage trays). As prevalence becomes very high, the critical region begins to increase in size again. This accounts for a switch to more situations of higher perceived threat combined with low to middling perceived efficacy of the health intervention.

**Figure 4:**
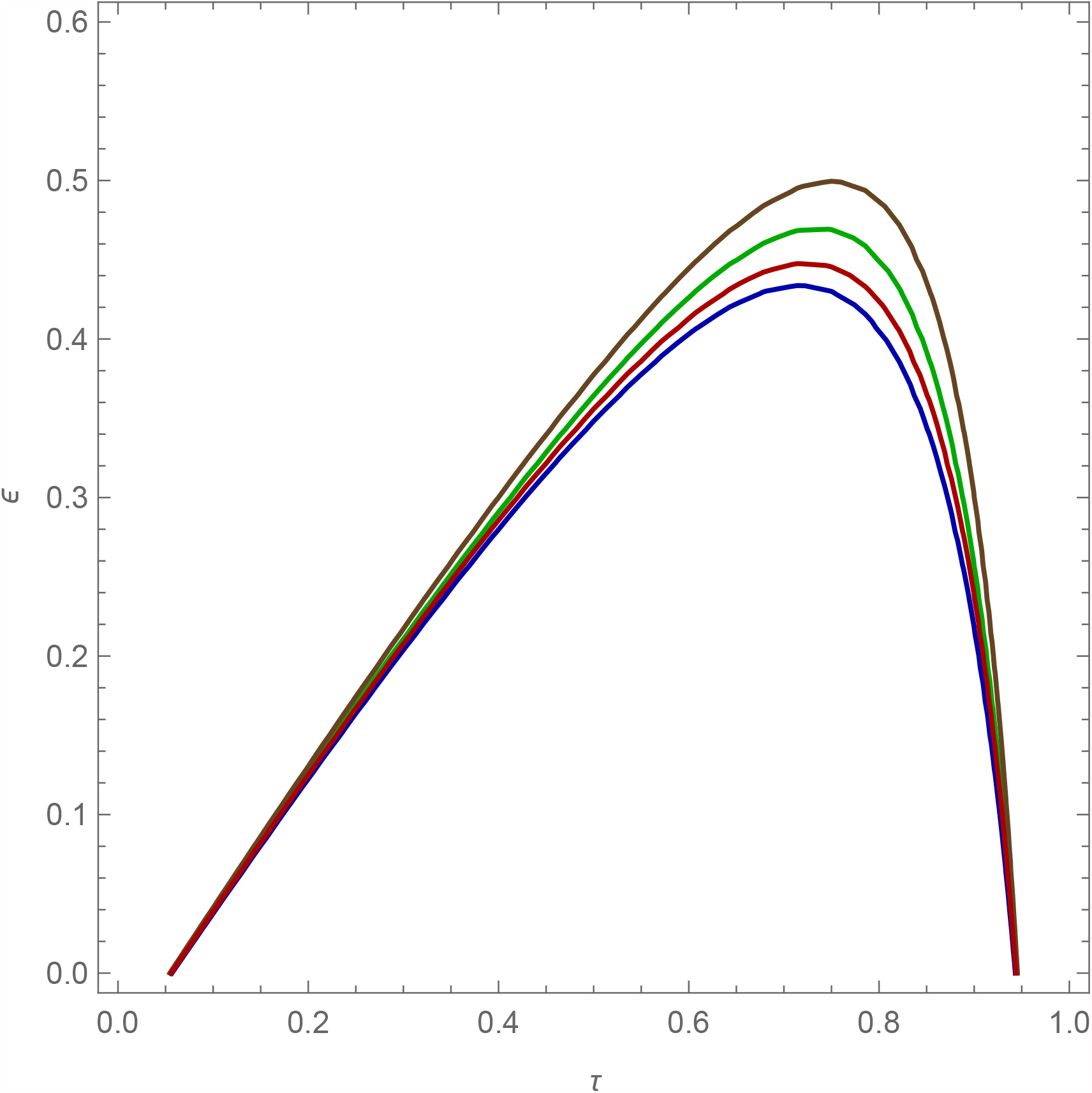
Contour plot show boundary in threat-efficacy (*τ, ϵ*) space where the optimal solution changes for different values of community prevalence (*π*). Note that the feasible region is bounded below the line *τ* = *ϵ*, but this condition is not sufficient as stated in main text. Brown is *π* = 0.01, green is *π* = 0.1, blue is *π* = 0.5, red is *π* = 0.9

## 4 Discussion

We have presented a theoretical mathematical model which explicitly incorporates human behaviour in line with the behavioural theory EPPM, following the recommendation made by Weston et al. (2018). Placing hand sanitiser gels before and after luggage screening are the optimal response however, if there are constraints on the airport that mean an intervention can only be placed at one location, our probabilistic model showed optimal placement is affected by underlying assumptions about compliance behaviour. If compliance with using hand sanitiser gel after luggage screening is greater than or equal to compliance before luggage screening then it is always optimal to place the intervention after luggage screening. This result is intuitive if individuals act in a self-protective way as the risk of contamination comes from handling the trays, so the self-protective behaviour would be performed after handling the trays. However, if we assume that the risk of contamination from the trays leads to a fear response in the individual then they may not act in a adaptive way (Witte, 1992). That is, if they perceive the threat of contamination from the trays to be high and the recommended intervention of using hand sanitiser gel afterwards to be low then individuals may not comply with the intervention. In such a situation it may be optimal to place hand sanitiser gel prior to luggage screening, where people may comply with using the gel and reduce the chance of contaminating trays, in turn reducing the risk of trays contaminating other individuals.

We consider this model to be a proof of concept. Its value is in demonstrating how more complex human behaviours can be explicitly captured in infectious diseases models, drawing on expertise from both the behavioural sciences and infectious disease modelling work, both of which have contributed greatly to COVID-19 response in the UK (see SAGE). We can make no formal assertion about the size of the critical region of parameter space where hand sanitiser gel before luggage screening is optimal as we do not have data to inform either the disease transmission model parameters, nor results to enable us to quantify measures of perceived threat and perceived efficacy in the behavioural component of the model. Yet the existence of such a critical region is sufficient to warrant further study. Liu et al. (2016) argue that hand hygiene must be practised soon after contact with the fomite in order for it to have any effect in reducing transmission, so it is likely a very rare situation where placing hand sanitiser gel prior to luggage screening is likely to be optimal. Yet if that situation occurred, what would be required is effective messaging to avoid the inflation of perceived threat (thereby reducing the likelihood of threat overwhelming efficacy) and to emphasise the efficacy of hand sanitiser gel. Hand hygiene alone will not be an adequate control measure, requiring a complementary intervention for the surfaces (Hurst, 1996). In our scenario this would be the cleaning of the luggage trays which we do not currently capture in our model. Since the current COVID-19 pandemic, some consideration has been made regarding decontamination of airport trays (Cadnum et al., 2020) indicating that this is an area where improvements can be made from a global health safety perspective. While we have not found evidence from environmental surveys of airport trays, proxy information to populate the model may be available from other airport-based studies (Coleman et al., 2018; Memish et al., 2014; Bailey et al., 2018; Nasir et al., 2016), other types of airports such as underground trains (Simon-Lewis, 2016), or other settings where a variable population interacts with a potentially contaminated object, such as supermarket trolleys and baskets (Mizumachi et al., 2011; Carrascosa et al., 2019). Likewise, we could obtain actual compliance data by observing use of the hand hygiene intervention in the airport but this would only be meaningful during a disease outbreak to elicit actual compliance when operational constraints and ethical considerations would be present.

Studies are not universally in agreement on the effectiveness of hand sanitiser gel on removing pathogens (for example, see Turner et al. (2010); Hovi et al. (2017); Savolainen-Kopra et al. (2012). Currently we do not capture this potential variation within our model as we assume hand sanitiser, when used, completely removes any pathogen. We could introduce variation in intervention effectiveness by multiplying the compliance probabilities, *ρ* and *ω*, by an additional effectiveness parameter. In the absence of data, we have also made the simplifying assumption that compliance would be the same for all individuals, however many factors could affect the chance of an individual complying with recommended health-protective behaviour. Factors could be individual characteristics of the traveller such as their age, or could be related to their reason for travel (business, or social) and who they are traveling with (alone, with family, with friends). Self-protective behaviour within the airport itself is not commonly examined; indeed, a screening of 1371 titles and abstracts accessed via PsychINFO, EMBASE, Medline and PubMed on 6th and 7th November 2017 revealed only 15 papers related to health protective behaviours in airports. From this literature we learnt that being male, social travel, last minute travel, and being younger, or at least not being in the mid-range of age, is more closely associated with not following health protective behaviour and advice. If sufficient data were available to inform our model then we could adapt it to allow for a population stratified by sex, age, and travel reason, with each group having different expected compliance.

## Data Availability

no data

## Funding

IH, CW, and MBND’s contribution was funded by Prevention and Management of High Threat Pathogen Incidents in Transport Hubs (PANDHUB, EU grant agreement number 607433), DW and IH are affiliated to the National Institute for Health Research Health Protection Research Unit (NIHR HPRU) in Modelling Methodology at Imperial College London. IH is supported by the National Institute for Health Research Policy Research Programme in Operational Research (OPERA, PR-R17-0916-21001), by Alan Turing Institute for Data Science and Artificial Intelligence, EPSRC (EP/V027468/1) and by UKRI through the JUNIPER modelling consortium (grant number MR/V038613/1).

## Acknowledgements

Thank you to JRK and JMT for helpful comments on earlier drafts. The views expressed are those of the authors and not necessarily those of PANDHUB, Public Health England, the National Health Service, the NIHR, the Department of Health and Social Care, Imperial College London or University of Manchester.

## Notes

### Competing Interest Statement

The authors have declared no competing interest.

